# Functional Characterization of Lipid Regulatory Effects of Three Genes Using Knockout Mouse Models

**DOI:** 10.1101/2021.07.01.21259304

**Authors:** Chen Yao, Holly Savage, Tong Hao, Gha Young Lee, Yuka Takemon, Wenting Bian, David E Hill, Marc Vidal, Ron Korstanje, Daniel Levy

## Abstract

Integrative analysis that combines genome-wide association data with expression quantitative trait analysis and network representation may illuminate causal relationships between genes and diseases. To identify causal lipid genes, we utilized genotype, gene expression, protein-protein interaction networks, and phenotype data from 5,257 Framingham Heart Study participants and performed Mendelian randomization to investigate possible mechanistic explanations for observed associations. We selected three putatively causal candidate genes (*ABCA6, ALDH2*, and *SIDT2*) for lipid traits (LDL cholesterol, HDL cholesterol and triglycerides) in humans and conducted mouse knockout studies for each gene to confirm its causal effect on the corresponding lipid trait. We conducted the RNA-seq from mouse livers to explore transcriptome-wide alterations after knocking out the target genes. Our work builds upon a lipid-related gene network and expands upon it by including protein-protein interactions. These resources, along with the innovative combination of emerging analytical techniques, provide a groundwork upon which future studies can be designed to more fully understand genetic contributions to cardiovascular diseases.

## Introduction

Cardiovascular disease (CVD) remains the leading cause of death worldwide^1^. A key goal of CVD research is the identification of specific genes and genetic variants that contribute to the disease^2^. Genome-wide association studies (GWAS) have revealed numerous inherited DNA sequence variants associated with CVD and its risk factors^3-5^. For many known CVD-associated variants, however, the biological mechanisms and underlying causal genes remain largely unknown. The overwhelming majority of CVD-related genetic variants lie within intergenic or noncoding regions of the genome ^3^, indicating that these single nucleotide polymorphisms (SNPs) impact CVD risk indirectly, for example, by altering transcription levels of nearby (*cis*) or remote (*trans*) genes. The identification of genetic variants that alter gene expression – expression quantitative trait loci (eQTLs) – has paved the way for functional studies linking candidate genes to disease^6^.

We postulated *a priori* that a network approach could portray biological systems underlying CVD traits and provide insights into disease-related genes and pathways. Network-based approaches have many biological and clinical applications and have yielded promising results in recent studies^7, 8^. Not only do networks allow for the integration of different types of biological information, but they also allow for unbiased discovery of disease genes that, when integrated with protein-protein interactions (PPI), can provide mechanistic explanations for diseases^9^. In a previous study, we linked 21 CVD traits based on their shared SNP associations, and in doing so, recapitulated the clustering of metabolic risk factors observed in epidemiological studies ^10^. Here, we expand that network model to identify causal genes and pathways for lipid traits and tested three genes for causality.

By integrating high-quality PPI data^11^ with SNPs associated in GWAS of lipids, gene expression, and fasting blood lipid levels in 5,257 Framingham Heart Study (FHS) participants (Supplementary table 1), we selected *ABCA6, ALDH2*, and *SIDT2* as candidate causal genes for lipid traits (low-density lipoprotein (LDL) cholesterol, high-density lipoprotein (HDL) cholesterol, and triglycerides (TG)). Dysregulation of circulating lipid levels has been implicated in the pathogenesis of CVD^12^. To confirm that the candidate genes identified from the network model are causal for the corresponding lipid traits, we created mouse knockout (KO) models to assess, *in vivo*, the consequences of perturbing key genes as well as the networks in which they function. The mouse KO models were therefore phenotyped for the corresponding lipid traits and additionally liver gene expression was measured by RNA-Seq to study perturbations within the network (Figure 1).

**Figure 1.**
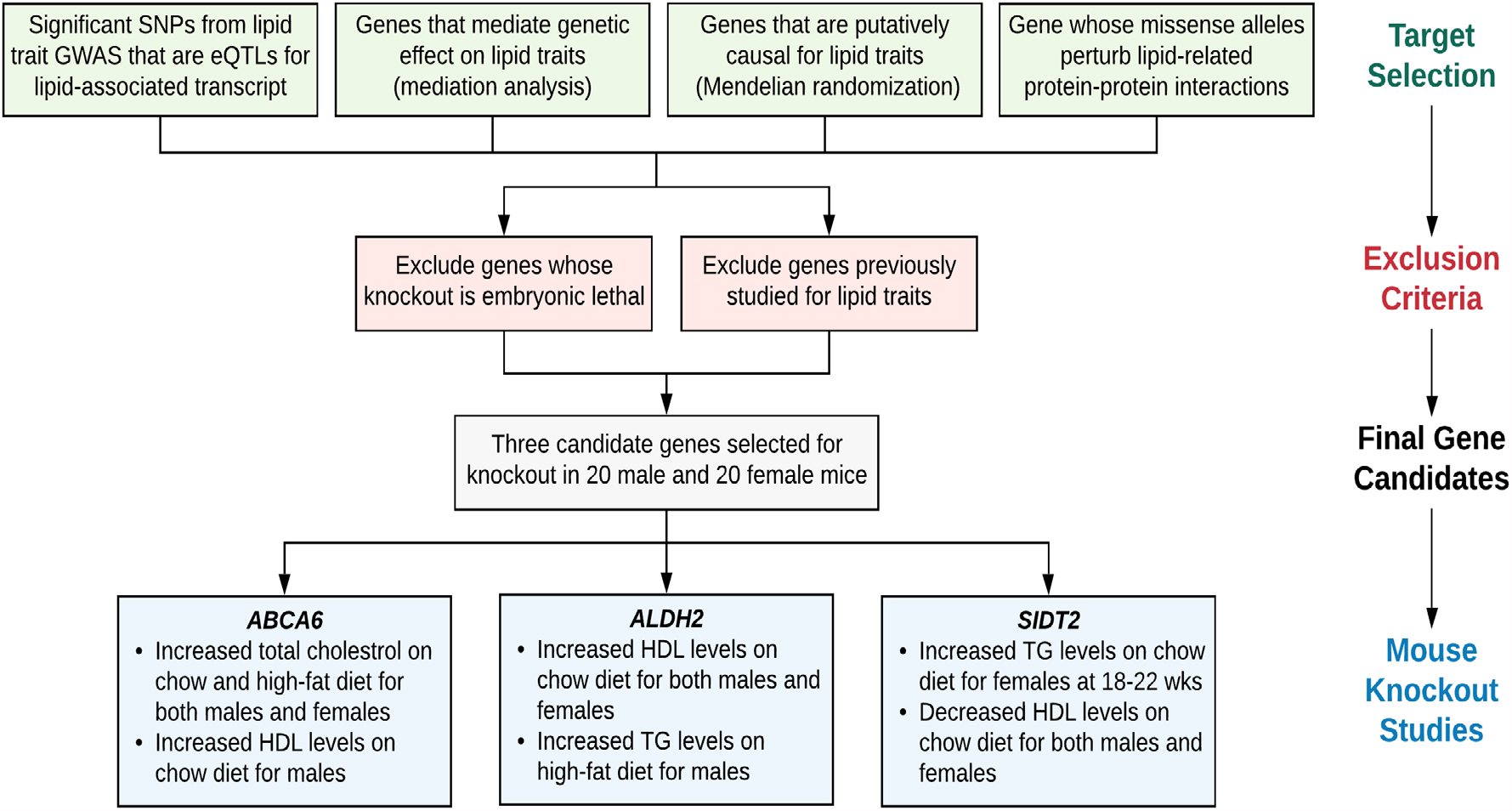
Study Design. Schematic diagram of stepwise approach to identify target genes by utilizing human lipid GWAS datasets, gene expression, protein interaction network and phenotype data, cross-reference with mouse knockout database, prioritize genes, and validate candidate genes.

We posit that creating a network based on the relationships among genes, traits, and PPIs can further our understanding of the genetic basis of lipid traits and their contributions to lipid dysregulation. By combining human genotype data with mouse KO experiments, we also provide a framework for selecting and validating lipid-related candidate genes.

## Results

### Identifying lipid trait-related expression quantitative loci

We identified 4670 (84%) lipid trait-associated expression quantitative loci (eQTLs; SNPs association with gene expression) by linking 5568 lipid trait GWAS SNPs (Supplementary Table 2) that were also associated with gene expression in whole blood in 5,257 Framingham participants^13^ (Supplementary Table 3). Seventy-two percent of these eQTLs affected the expression of a nearby (*cis*) transcript (SNP located within 1 Mb of the transcript), suggesting that they may play a role in regulating gene expression. For these eQTLs, we conducted mediation testing to detect if the association of the GWAS SNP with the corresponding trait was mediated by gene expression. At P<0.005, we identified 464 SNPs with significant mediation effects on lipids traits (Supplementary Table 4).

### Expanding the candidate gene network

To expand the candidate gene network, we built a CVD gene network by incorporating all protein coding genes containing CVD GWAS SNPs. Our CVD-centric network contained binary PPIs for 846 CVD-associated proteins (out of ∼1,300 tested) extracted from a dataset of ∼58,000 binary PPIs among 10,690 human proteins obtained from both systematic binary mapping and literature curations^14^. The 846 CVD-associated proteins and their first-degree neighbors constituted a network of ∼8,600 interactions among 4,336 proteins (Supplementary Table 5), including 444 intra-CVD PPIs among 349 lipids-associated proteins (Supplementary Table 6). This expanded network is amenable to investigating disease models by analyzing other features such as shared gene ontology (GO) terms, shared expression profiles, and functional similarity.

#### Ranking candidate genes

Combining GWAS lipid trait SNPs, eQTLs, and mediation results, we selected seven candidate genes to test against three lipid traits in Mendelian randomization (MR) analyses: *DOCK7, TAGLN, SIDT2, ALDH2, SLC44A4, ABCA6*, and *ATG4C* (See Methods for gene selection). We used genetic variants (focusing on those that are eQTLs variants) as instruments to investigate the causal relations between gene expression and lipid phenotypes. The causal effects of the seven genes were further tested by MR using independent GWAS data from the Global Lipids Genetics Consortium (GLGL)^15^ (Table 1) and filtered based on 1) gene novelty (not previously studied in a mouse model for cardiovascular phenotypes), 2) repository availability of knockout sperm or embryos that have been shown to produce viable animals, and 3) interaction with CVD-related proteins in an integrated network based on PPIs. This approach identified three genes as suitable for detailed mouse KO experiments: *ABCA6* for non-HDL, *ALDH2* for HDL, and *SIDT2* for TG (Table 2). Though *ABCA6* was not found to be causally associated with lipids traits by MR. rs740516, a variant in *ABCA6*, was significantly associated with LDL in GWAS (P=7×10^−9^) and was associated with expression of *ABCA6* (P=3.3×10^−5^). *SIDT2* has been previously studied in relation to lipid metabolism and homeostasis, our finding is one of the first to utilize an integrative genomics framework to better understand the genetic basis of its effect on lipid traits^16, 17^. The effects of these genes on secondary lipid traits (*ABCA6* on HDL and TG, *ALDH2* on non-HDL and TG, *SIDT2* on HDL and non-HDL) are also summarized.

**Table 1.**
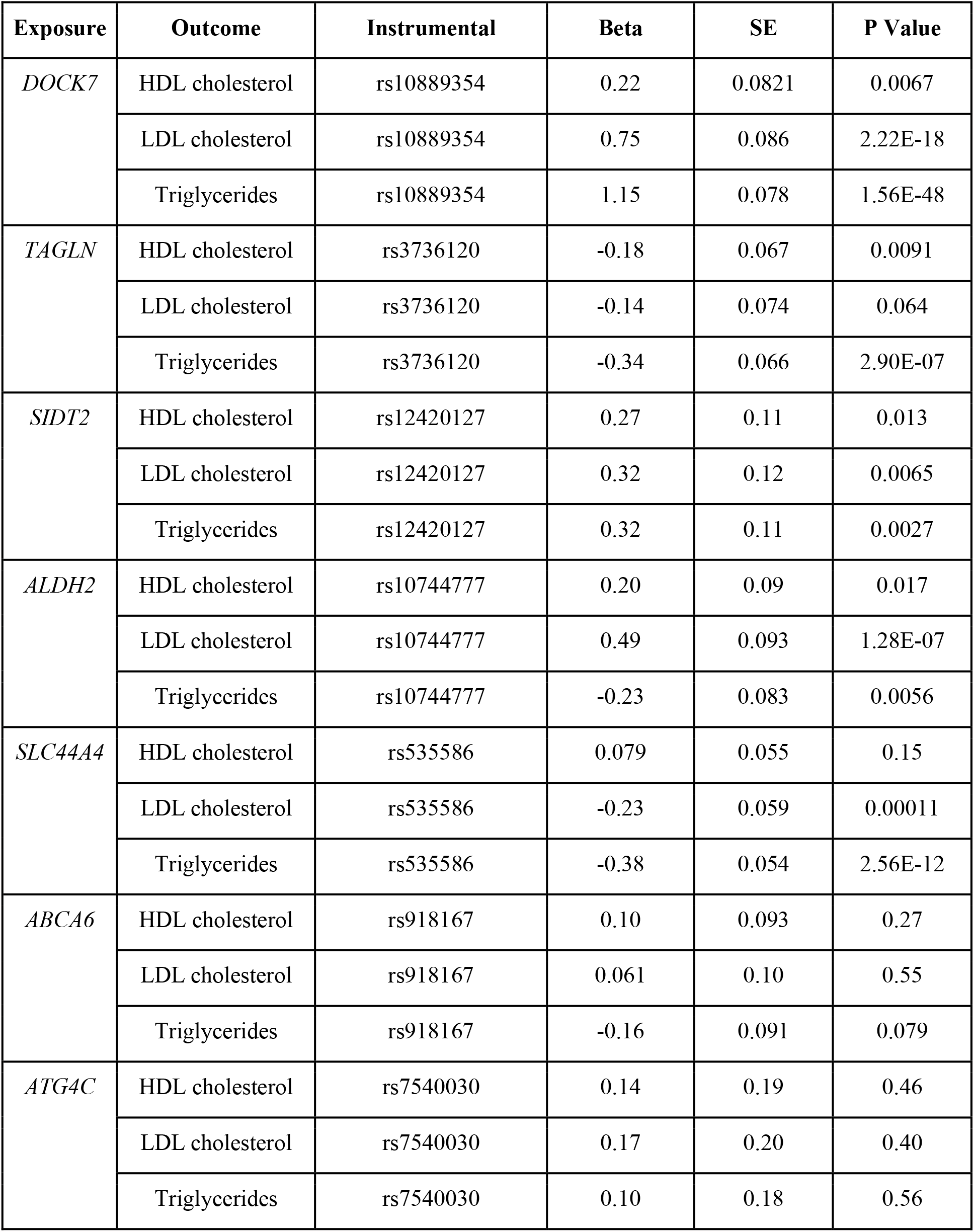
MR tests for candidate genes.

**Table 2.**
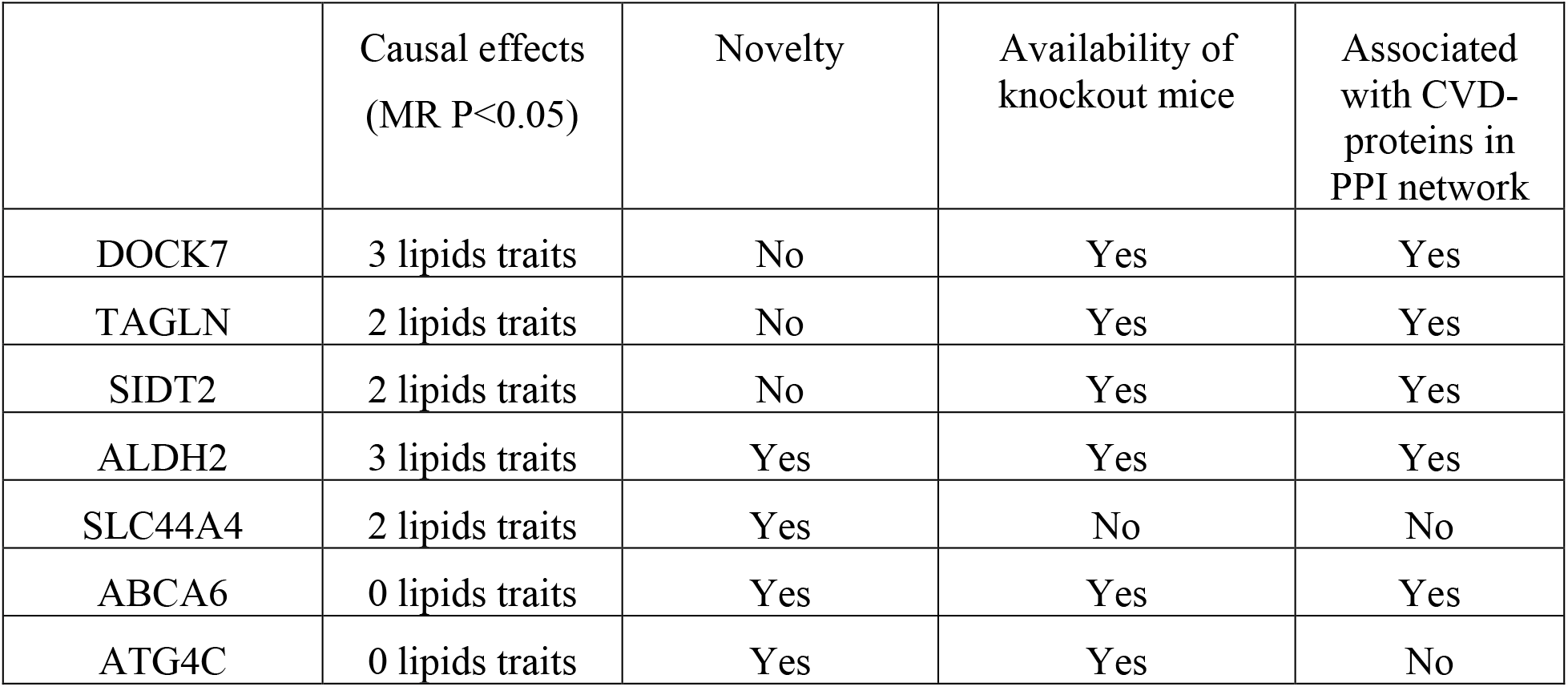
Criteria of candidate gene selection.

### *ABCA6, ALDH2, SIDT2*, and the corresponding mouse knockout models

#### *ABCA6* Knockout

The protein ABCA6, encoded by *ABCA6*, is a member of the superfamily of ATP-binding cassette (ABC) transporters. ABC proteins transport various substrates, including lipids, peptides, vitamins, and ions^18^, across extra- and intracellular membranes. *ABCA6* is expressed exclusively in multicellular eukaryotes and has been suspected of acting as an intracellular transporter in macrophage lipid homeostasis^19^.

Comparing total cholesterol and non-HDL cholesterol levels (which are primarily comprised of LDL cholesterol) between C57BL/6N-*Abca6*^*tm2a(KOMP)Wtsi*^/TcpRkorJ mice (*Abca6* knockout) and WT C57BL/6N mice, we observed a significant increase in total cholesterol in males on the chow diet at 22 weeks of age and in males on the high-fat diet at both 18 and 22 weeks of age (Figure 2). A similar trend was identified in females.

**Figure 2.**
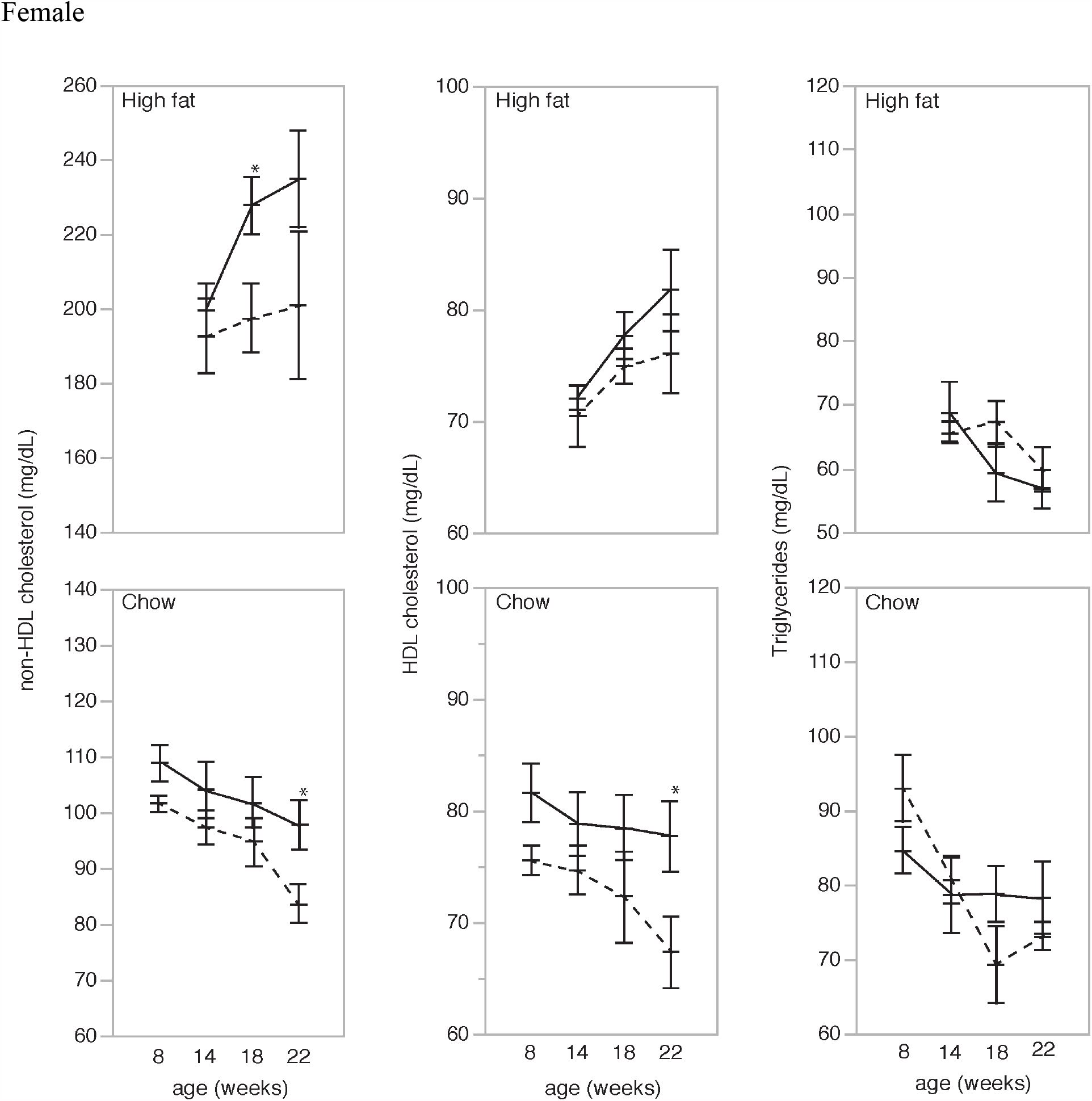
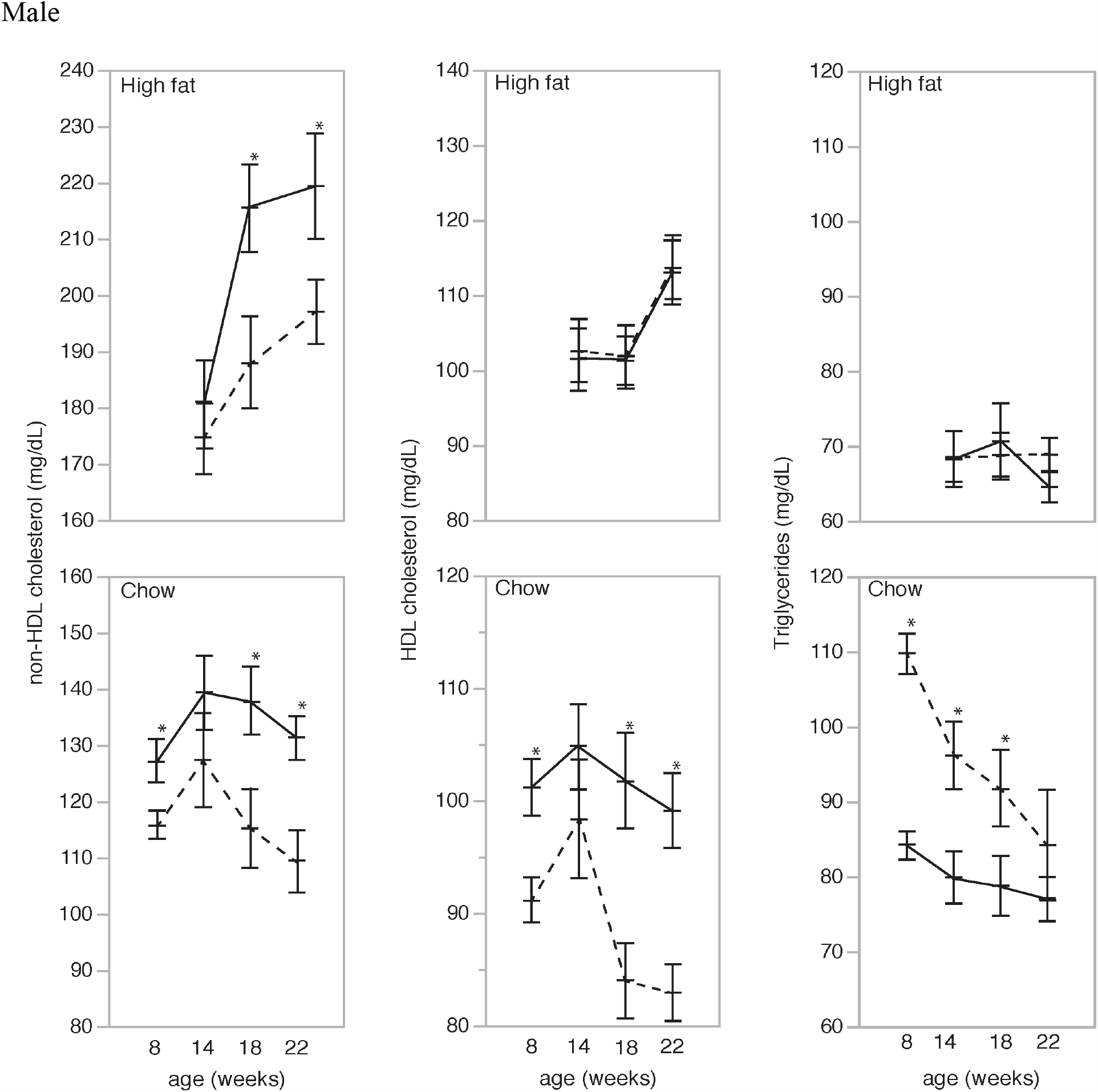
Comparison between *Abca6* knockout (solid line) and wildtype (dotted line) mice in females and males on regular chow and high-fat diet. Ten mice for each genotype and sex per diet group. Asterisk indicates a significant difference (P<0.05) between knockout and wildtype at the specific time point.

In addition, knockout of *ABCA6* led to a significant increase in HDL cholesterol in male mice on the chow diet but not on a high-fat diet with a similar but nonsignificant trend observed in females. This finding is supported by previous studies that have shown that *ABCA6* is involved in reverse cholesterol transport, specifically by facilitating phospholipid and cholesterol export from the cell^19, 20^. There were no significant differences in TG levels in the female mice on both chow and high-fat diet, but the male *Abca6*^-/-^ mice had lower TG levels on the chow diet.

#### *ALDH2* Knockout

*ALDH2* encodes aldehyde dehydrogenase, the second enzyme of the major oxidative pathway of alcohol metabolism. This gene encodes a mitochondrial isoform, which has a high affinity for acetaldehydes and is localized in the mitochondrial matrix. Deficiencies in *ALDH2* have been shown to be correlated with differences in lipid levels due to its role in the metabolism of lipid-peroxidation-derived aldehydes ^21^.

*ALDH2* was causally related to all three lipids in MR (Table 1). In addition, SNPs in *ALDH2* were significantly associated with HDL and LDL in GWAS and gene expression in FHS (Supplementary Table 3). Comparing HDL cholesterol levels between B6Dnk;B6N-*Aldh2*^*tm1a(EUCOMM)Wtsi*^/IegRkorJ mice (*Aldh2* knockout) and WT C57BL/6N mice, we found significantly increased HDL cholesterol levels in both female and male *Aldh2* KO mice at several time points compared to controls when fed the chow diet (Figure 3). On a high-fat diet, we no longer observed a difference in males, but there was a significant difference in females at 18 weeks of age.

**Figure 3.**
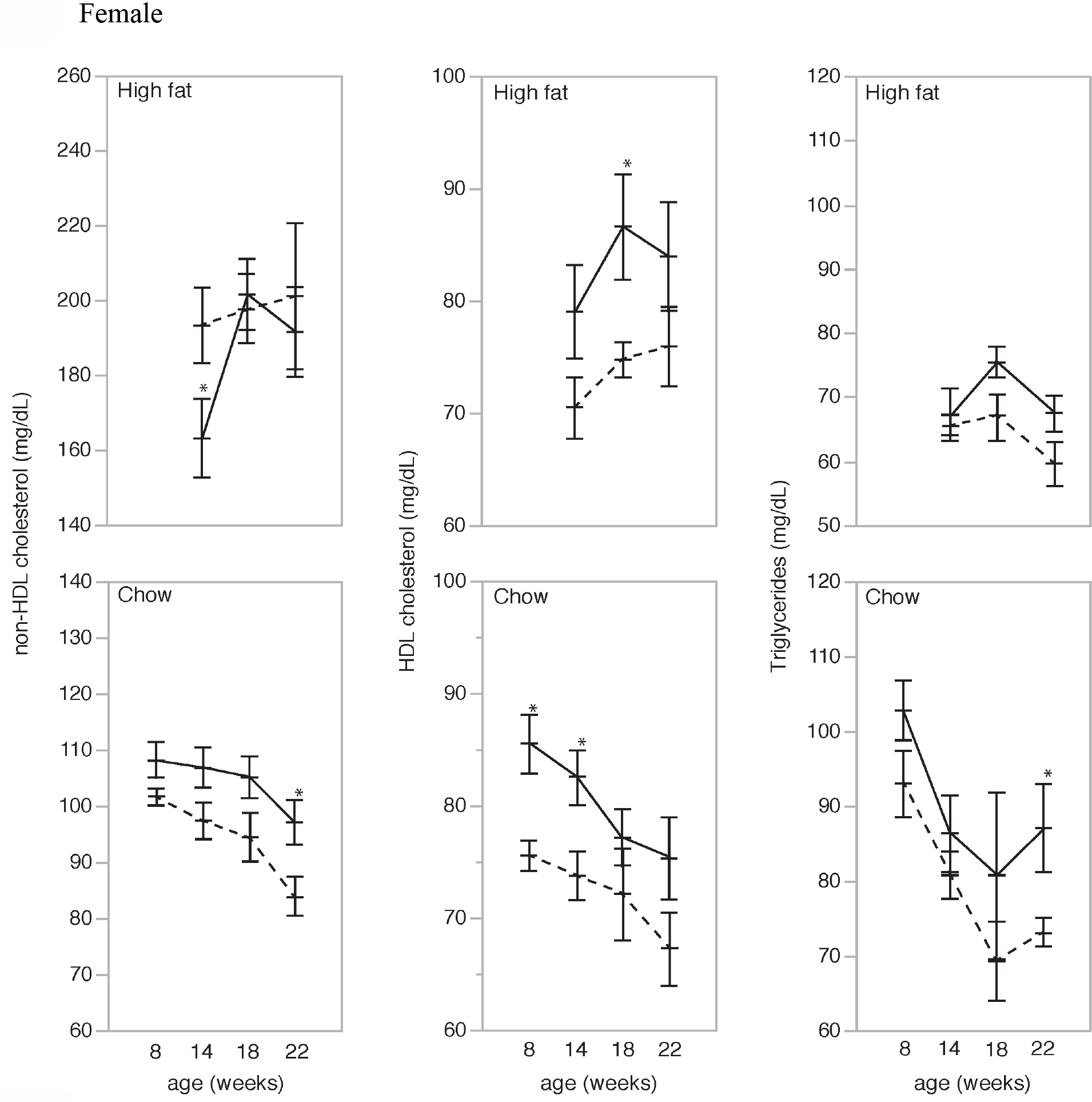
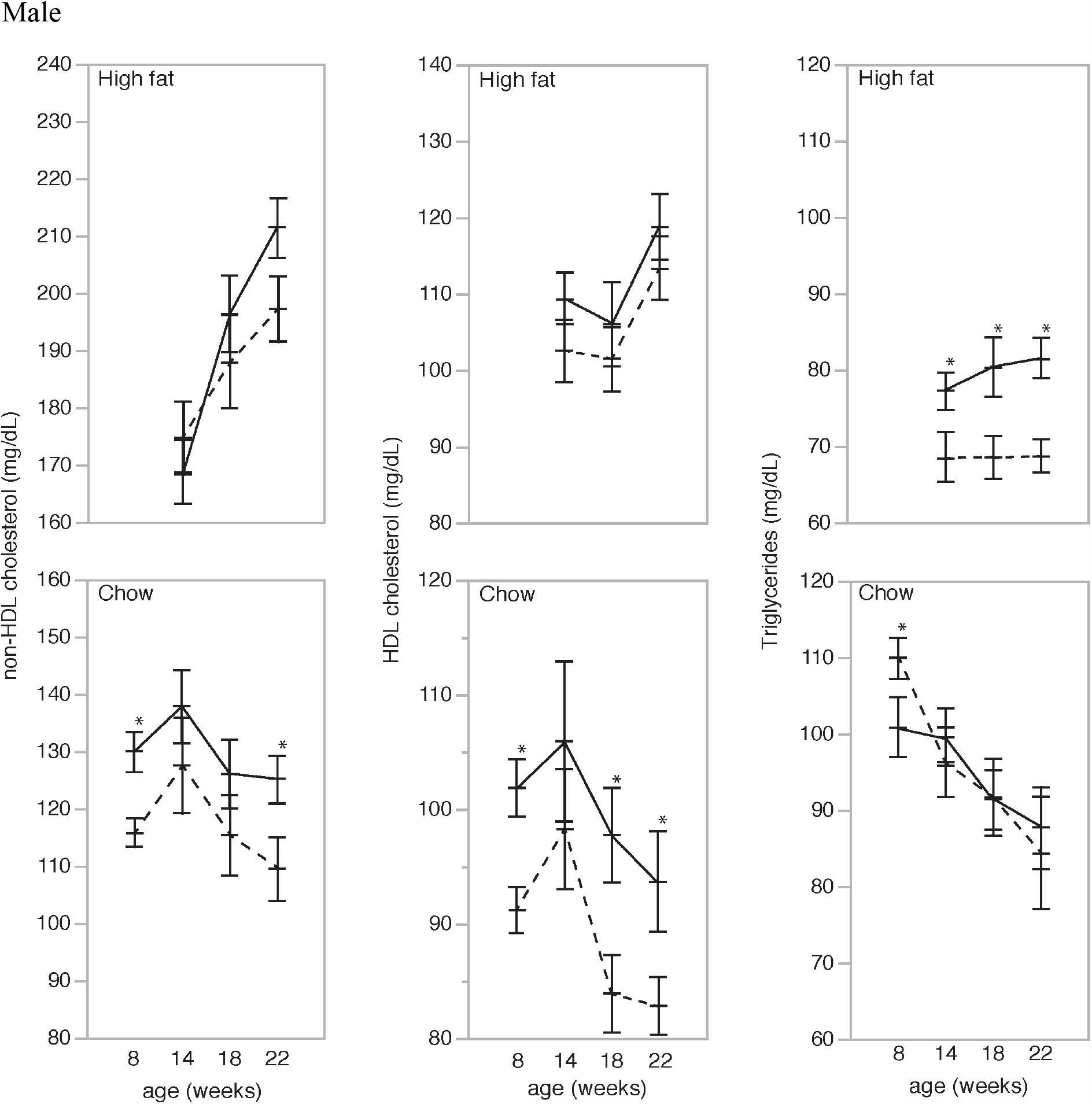
Comparison between *Aldh2* knockout (solid line) and wildtype (dotted line) mice in females and males on regular chow and high-fat diet. Ten mice for each genotype and sex per diet group. Asterisk indicates a significant difference (P<0.05) between knockout and wildtype at the specific time point.

Except for a few specific time points, there were no differences in non-HDL levels for either diet in males or females. There were no differences in TG levels in the female mice, but in the male mice we observed increased TG levels in the KO animals on a high-fat diet. This suggests that the knockout of *Aldh2* in males might lead to a decreased risk of CVD on the chow diet, but an increased risk on the high-fat diet.

#### SIDT2 Knockout

SIDT2 is a transmembrane protein that predominantly localizes on lysosomes but is also detectable in the plasma membrane of human embryonic kidney cells. Overexpression of SIDT2 in some cells is accompanied by a significant reduction of detectable lysosomes, indicating that the overexpressed protein leads to lysosomal dysfunction. Lysosomes are thought to be the major intracellular compartment for the degradation of macromolecules and play an important role in regulating lipid degradation pathways^22, 23^. Previous *SIDT2*-knockout experimentation has shown that SIDT2 plays a major role in regulating lipid autophagy and metabolism and cholesterol and triglyceride transport in mammalian cells, particularly in the liver^16, 17^.

*SIDT2* was significant in MR test for all three lipids traits. SNPs in *SIDT2* were significantly associated with LDL in GWAS and gene expression in FHS (Supplementary Table 3). We identified *SIDT2* in our human cohort to be associated primarily with triglycerides and therefore compared TG levels between B6;129S5-*Sidt2*^*tm1Lex*^/MmucdRkorJ mice (*Sidt2* knockout) and WT control littermates. We observed a significant increase in TG levels in the female mice on the chow diet at 18 and 22 weeks of age (Figure 4). Male TG levels were only increased at 8 weeks of age.

**Figure 4.**
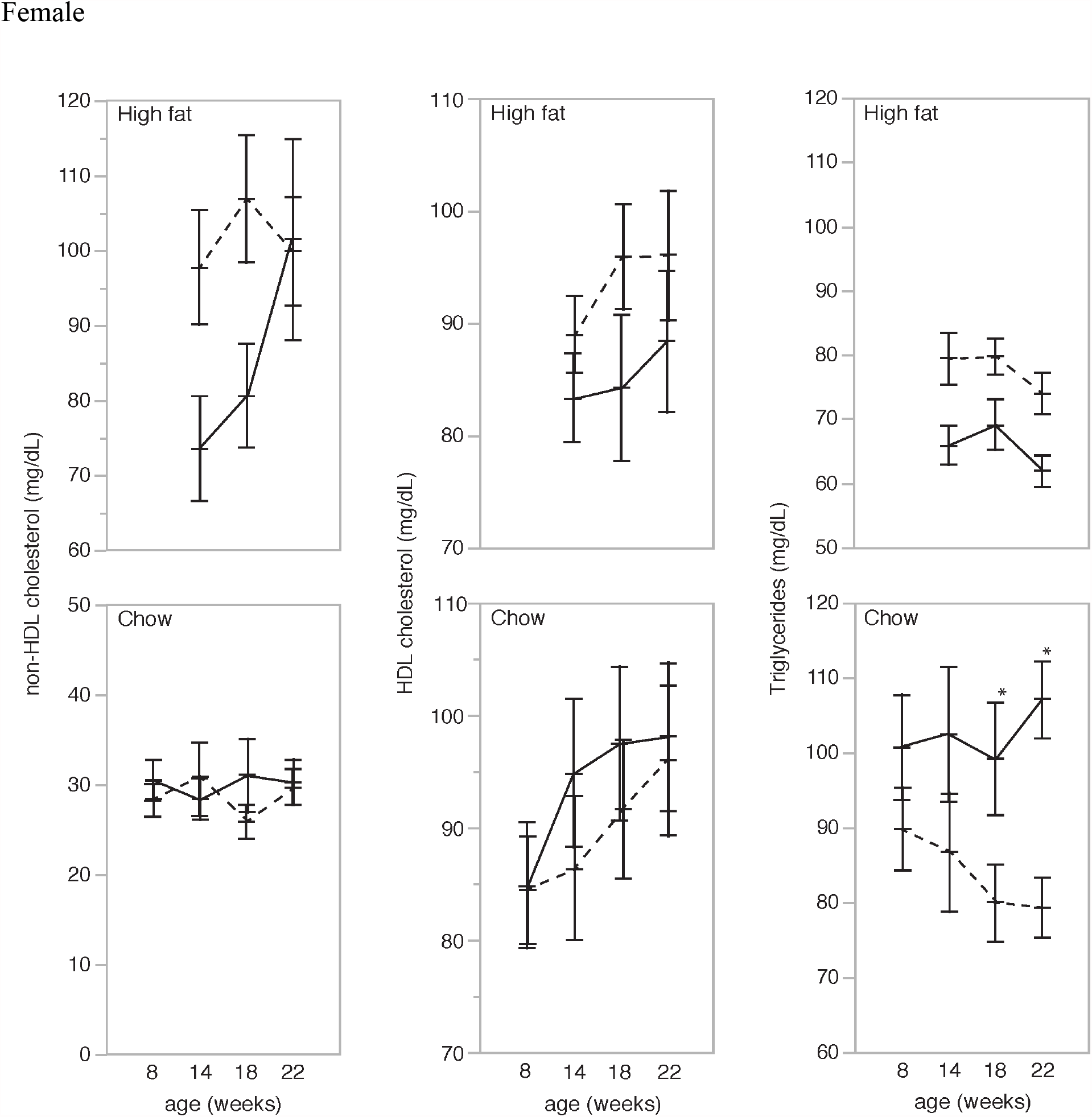
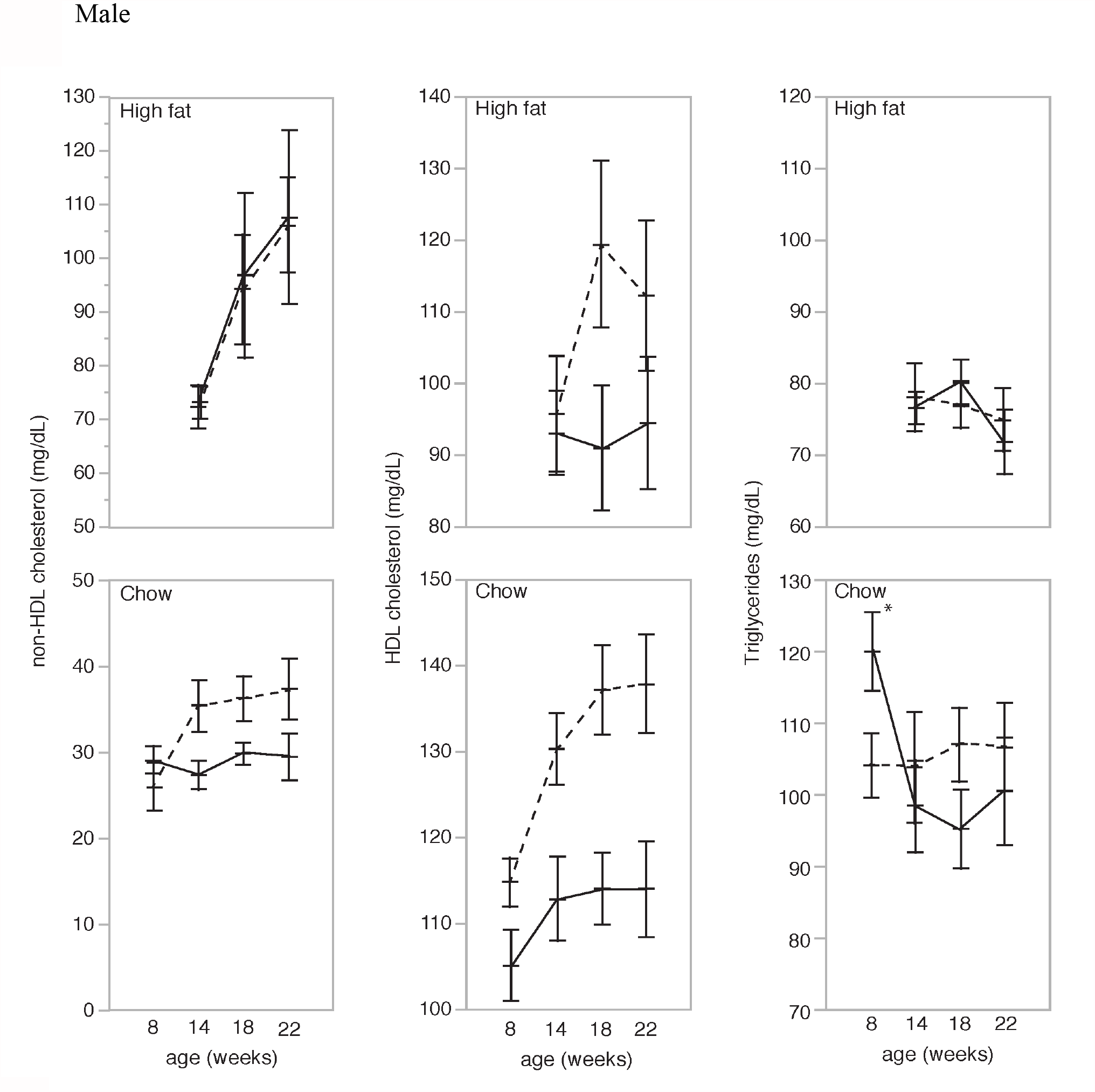
Comparison between *Sidt2* knockout (solid line) and wildtype (dotted line) mice in males and females on regular chow and high-fat diet. Ten mice for each genotype and sex per diet group. Asterisk indicates a significant difference (P<0.05) between knockout and wildtype at the specific time point.

In addition, HDL cholesterol levels were significantly decreased in male and female KO mice on the high-fat diet. On a chow diet, HDL cholesterol levels appeared to decrease even further in male mice. HDL cholesterol levels, however, increased slightly in females. Female non-HDL cholesterol levels did not change on the chow diet; in the early stages there was a large decrease in non-HDL cholesterol on the high-fat diet that ultimately dissipated. Male non-HDL cholesterol levels did not change on the high-fat diet and significantly increased on a chow diet.

### Expression changes after gene knock out

To explore transcriptome-wide alterations resulting from knocking out the target genes, we conducted RNA-Seq for *ALDH2* and *ABCA6* in three male and three female knockout animals and three male and three female control littermates for each of the two diets. We didn’t conduct the RNA-Seq for *SIDT2* in light of previously reported studies of this knockout ^16, 17^. At FDR<0.05 (T test, P<0.014), we identified an average of 238 differentially-expressed genes (ranging from 69 to 403 for eight comparisons) after knockout (Supplementary Table 7). The differentially-expressed genes were analyzed through the use of Ingenuity Pathway Analysis (IPA,QIAGEN Inc.,)^24^. Function enrichment analyses were conducted on the differentially expressed genes using curated information from the QIAGEN Knowledge Base^24^. Though canonical pathways of differentially expressed genes from mice of different diet conditions are enriched in different metabolic and cell signaling pathways, the affected diseases and molecular functions are highly consistent with regard to metabolic disease and lipid metabolism (Supplementary figure 1). The top toxicological functions were liver steatosis and cardiac dysfunction. Network analysis based on Ingenuity Knowledge Base^24^ revealed that *Abca6* affected cholesterol level via cytokine IFNG (interferon gamma) and that *Aldh2* affected cholesterol level through enzyme GNMT (glycine N-methyltransferase) and acetaldehyde (Supplementary Figure 2). Comparing RNA-seq data from *Abca6* and *Aldh2* knockout of the same sex and diet, we found 19%-60% overlap of differentially expressed genes. Using IPA functional enrichment at P<0.005, we found that the overlapping genes are enriched in lipid metabolism (Supplementary Table 8), suggesting a common pathway affected by these two very different genes.

## Discussion

To represent the complex molecular dynamics that underlie CVD, we constructed a network that integrates SNPs, gene expression, protein-protein interactions, and lipid phenotypes. From this integrated network approach, we identified three genes – *ABCA6, ALDH2*, and *SIDT2* – that were linked to lipid levels and were investigated further through mouse knockout models. The mouse experiments recapitulated network-predicted relations of all three genes to the corresponding lipid traits and provided experimental support for the integrative approach we developed.

Disease processes involve many interacting molecules and physical and biochemical processes, and thus the analysis of single data types is often insufficient to explain the etiology of complex traits. For example, carriers of risk alleles do not always have phenotypic consequences as such genetic variants do not necessarily alter the expression of disease-related genes or proteins. Therefore, in order to draw a more comprehensive view of biological processes, high-throughput data from different elements of multidimensional molecular information must be integrated and analyzed.

Results from our mouse knockout models provide proof of principle for the utility of the integrative network approach that we developed. Results of the mouse experiments largely support our network model, with a few exceptions. Knockout of *Abca6*, which contains GWAS SNPs for LDL cholesterol and total cholesterol, resulted in higher total cholesterol (which is largely LDL cholesterol) and HDL cholesterol in male mice, suggesting a potential role of this gene in CVD development. *ABCA6* plays a key role in macrophage lipid homeostasis by facilitating phospholipid and cholesterol export from the cell^19, 20^. *ABCA6* dysfunction may cause a buildup of intracellular phospholipids and cholesterol. Knockout of *Aldh2*, which was chosen for its putatively causal association with HDL cholesterol, resulted in significantly increased HDL cholesterol levels in both female and male mice. Aldehyde dehydrogenase, the protein encoded by *ALDH2*, is an enzyme in the oxidative pathway and plays a major role the metabolism of lipid peroxidation-derived aldehydes ^21^. In our knockout models, the absence of *Aldh2* resulted in higher lipid levels, which supports the idea that *ALDH2* is involved in lipid degradation/clearance. Knockout of *Sidt2*, which contains GWAS SNPs for HDL cholesterol, resulted in a significant increase in triglyceride levels in the females on the chow diet at 18 and 22 weeks of age. SIDT2 plays a crucial role in the uptake and intracellular transport of triglycerides and cholesterol ^22, 23^. Previous studies^16^ have shown that the absence of *Sidt2* causes increased serum triglycerides and free fatty acids in mice. After analyzing the knockout results, we compared them with expression data for the corresponding human genes. We discovered that while each gene knockout produced some significantly different results in the female and male mice, FHS human gene expression data did not reveal sex differences in lipid effects for any of the genes (P>0.05, data not shown). This may be due to innate differences in lipid metabolism and regulation between the two species.

In contrast to similar studies that have relied on public databases to identify eQTLs^25^, our study takes a direct and thorough approach, analyzing extensive phenotypic datasets in relation to gene expression in human peripheral blood. This method, combined with mediation analysis, allowed us not only to identify significant pathway enrichment from GWAS SNPs, but also to investigate mechanisms underlying lipids traits. Our study is also unique in that we combined several emerging techniques to identify and validate genes; these techniques included the use of functional eQTL studies and the integration of PPI data. Functional studies, especially those involving the use of networks to mirror biological systems, have shown great promise in the ability to identify candidate genes for various diseases^26^. In one study, Atanasovska et. al used a network-based approach to prioritize genes for hundreds of cardiometabolic SNPs to identify disease-predisposing genes^27^. The integration of PPI with eQTL analyses also has great potential to offer mechanistic explanations for diseases^11, 14^.

One limitation of our study is the use of whole blood for expression profiling. Although the lipid traits were measured in blood, and as such, whole blood-derived eQTLs may be highly relevant to the phenotypes we studied, in some cases, we observed opposite direction between MR predicted effects and mouse knockout (*ALDH2* on HDL). Another limitation is that our MR analyses could not infer sex-specific causal effects because the underlying GWAS studies were conducted in pooled-sex analyses, even though our mouse experiments revealed notable sex differences. Differential lipid metabolic responses between male and female mice have also been previously identified in other gene knockout studies^28^.

In summary, our network approach allowed us to identify novel candidate genes that may contribute to CVD via lipid effects. As such, these genes represent attractive targets for the treatment of dyslipidemia and the prevention of CVD. Looking forward, we plan to continue to expand the CVD network and investigate additional causal genes. As we do so, we anticipate that it will increasingly explain genes, pathways, and mechanisms underlying CVD and point toward promising precision drug targets. Our integrative network also provides insights into how dysfunction of novel CVD-associate genes is manifested at the molecular level ^29^. Approaches similar to those used in this study may not only be expanded upon in relation to CVD but also be used to investigate other diseases impacted by genetic and epigenetic factors. To make future studies more comprehensive and accurate, it will be important to expand eQTL databases to other disease-related tissues and to include additional clinically important traits as well.

### Materials and Methods Samples and Phenotypes

In 1948, the FHS started recruiting participants (original cohort) from Framingham, MA to begin the first round of extensive physical examinations and lifestyle surveys to investigate CVD and its risk factors ^30^. In 1971 and 2002, the FHS recruited offspring (and their spouses) and adult grandchildren of the original cohort participants into the offspring and third-generation cohorts, respectively ^31-33^. Plasma total cholesterol, HDL cholesterol, triglycerides, and glucose were measured in the morning after an eight-hour overnight fasting. Body mass index (BMI) was defined as weight (kilograms) divided by height squared (meters^2^). Current smokers were defined as those who smoked, on average, at least one cigarette per day during the year prior to the FHS clinical assessment. Clinical characteristics of the study sample are summarized in Supplementary Table 1. All participants from the FHS gave informed consent for participation in this study and for the collection of plasma and DNA for analysis. The FHS study protocol was approved by Boston Medical Center: Protocol ID: H-27984, Boston University Medical Center IRB (BUMC IRB) Title: FRAMINGHAM HEART STUDY BIOMARKER PROJECT.

#### Genotype data

5,568 SNPs that were associated with the three lipid traits – HDL cholesterol, LDL cholesterol, and triglycerides – at p≤5×10^−8^ (in the GRASP database, downloaded in June 2016, Supplementary Table 2) were curated and matched with the FHS 1000 Genomes Project imputed genotype data ^34^. SNPs with imputed quality score (r^2^) <0.3 and minor allele frequency (MAF) <0.01 were excluded, resulting in 4173 genome-wide significant SNPs for eQTL analysis.

#### Gene expression

Whole blood was collected in PAXgene™ tubes (PreAnalytiX, Hombrechtikon, Switzerland) and frozen at −80°C. RNA was extracted using a whole blood RNA System Kit (Qiagen, Venlo, Netherlands) and mRNA expression profiling was assessed using the Affymetrix Human Exon 1.0 ST GeneChip platform (Affymetrix Inc, Santa Clara, CA), which contains more than 5.5 million probes targeting the expression of 17,873 genes. The Robust Multi-array Average (RMA) package ^35^ was used to normalize the gene expression values and remove any technical or spurious background variation. Linear regression models were used to adjust for technical covariates (batch, first principal component, and residual mean of all probesets).

### Identification of LIPIDS-associated eQTLs

The eQTLs were identified from FHS genotype data and whole blood gene expression as described previously ^13^. eQTL analyses were conducted in two phases: 1) gene expression residuals were generated after accounting for the effects of sex, age, platelet count, white blood cell count, and imputed differential blood cell counts; analyses were performed using R version 3.0.1 with a mixed-effect modeling package that adjusted for familial relationships; and 2) underlying confounding factors were accounted for by 20 Probabilistic Estimation of Expression Residuals (PEERs)^36^ factors that were computed using the residualized expression data. The residualized expression was fit to a linear model using the PEER factors along with sex, age, and effect allele dosages. The algorithm was implemented with Graphical Processing Units (GPUs). *cis*-eQTLs were defined as SNPs that reside within 1 Mb up or downstream of the transcription start site. False discovery rate (FDR) computations for *cis*- and *trans*-eQTLs were computed separately. SNPs at FDR <0.05 were considered statistically significant eQTLs.

Putative causality was first tested by mediation tests, in which the underlying mechanism for a relationship between two variables is tested by introducing a third explanatory variable. The analysis was conducted with the Mediation Package^37^ in R where the “exposure” variable represented a SNP, the “mediator” or “explanatory variable” represented gene expression, and the “outcome” variable represented the phenotype. The mediation effect was measured on a scale from 0-100% where a 100% mediation effect indicated that the entire relationship between a SNP and a phenotype (direct effect) was explained by changes in gene expression, i.e., the “mediator” or the “explanatory variable.” Significant mediation effects were selected at a permutation p-value <0.005 (based on 1000 permutations).

Significant mediation effects were further tested by Mendelian randomization (MR)^38^ to determine causal associations between gene expression and phenotype. MR uses common genetic variants with well-understood effects on an exposure as instrumental variables (IV) to infer causality of an exposure to an outcome. This approach was applied to the aforementioned three lipid traits (LDL cholesterol, HDL cholesterol, and triglycerides). Here, the sentinel *cis*-eQTL (top eQTL located within 1 Mb of the tested gene), based on the lowest SNP-gene expression p-value from the 1000G GWAS, was selected as the IV for its corresponding gene in MR analysis. Based on the association between the sentinel *cis*-eQTL and the summary statistics from the three lipid GWAS of 188,577 individuals obtained from the Global Lipids Genetics Consortium (GLGL)^15^, the MR analysis was performed using the MR Base package^39^ and the two-sample MR method ^38^.

### Integration with PPI Networks

Binary PPIs were extracted from a systematically generated and literature-curated datasets, which in total contain ∼58,000 PPIs among 10,690 human proteins^11, 14^. Putatively causal variants from genotyping and gene expression analysis were integrated with PPI networks to assess the extent of potential network perturbations associated with disruption of PPIs or altered expression^9^. Network perturbation information on missense alleles for various lipids-associated genes, which assesses the degree to which a mutant protein exhibits an altered spectrum of PPIs relative to the WT protein and all other mutants^11, 14^, was also included. For proteins without direct (physical) interactions, we use predicted protein-protein interactions from the STRING database to expand the network^40^.

### Identifying Candidate Genes

All candidate genes were assessed by each of the each of the following criteria: 1) Does the candidate gene contain a SNP(s) associated with a lipid trait in GWAS (p<5×10^−8^) and is the GWAS SNP(s) is also an eQTL? 2) Is expression of the eQTL-associated gene also associated with the same lipid trait? 3) Are genetic effects on the lipid trait mediated by the expression of the eQTL-associated gene? 4) Does the gene test positive in MR (p<0.05)? 5) Have any interactions with CVD related proteins?

We then ranked the genes based on how many of the criteria were met and further excluded any genes if the knockout was known to be lethal based on existing literature. Three genes *ABCA6, ALDH2*, and *SIDT2* met more than three of these selection criteria and were selected for further investigation.

### Validation of Candidate Genes in Animal Models

To test the hypothesis that key genes identified in the LIPIDS networks can be validated in mouse models for the corresponding traits, we obtained KO mouse strains for the three genes identified as having causal effects on HDL cholesterol, LDL cholesterol, and triglycerides, respectively. C57BL/6N-*Abca6*^*tm2a(KOMP)Wtsi*^/TcpRkorJ, B6Dnk;B6N-*Aldh2*^*tm1a(EUCOMM)Wtsi*^/IegRkorJ, and B6;129S5-*Sidt2*^*tm1Lex*^/MmucdRkorJ KO mice were generated at The Jackson Laboratory using sperm or embryos provided by the International Knockout Mouse Consortium and maintained on a C57BL/6NJ genetic background. All animals were housed at The Jackson Laboratory, which is approved by the American Association for Accreditation of Laboratory Animal Care. Animals were kept on a 12-hour (6am-6pm) light/dark cycle with a room temperature between 68 and 72°F, and either fed a chow diet (5K52, LabDiet) or a high-fat diet (TD.06414, Teklad Custom Diet).

Cohorts of 20 males and 20 females of both KO strains and C57BL/6NJ control mice were raised on the chow diet. At 12 weeks of age, ten males and ten females from each cohort were switched to the high-fat diet while the other animals continued on the chow diet. Plasma samples were collected at 8, 14, 18, and 22 weeks after a four-hour fast. Total cholesterol, HDL cholesterol, and triglycerides were measured using a Beckman Coulter Synchron CX®5 Delta autoanalyzer. Non-HDL was calculated as the difference between total cholesterol and HDL cholesterol.

At 22 weeks of age, animals were euthanized and their livers were snap-frozen. RNA was isolated from liver tissue using the MagMAX mirVana Total RNA Isolation Kit (ThermoFisher) and the KingFisher Flex purification system (ThermoFisher). Tissues were lysed and homogenized in TRIzol Reagent (ThermoFisher). After the addition of chloroform, the RNA-containing aqueous layer was removed for RNA isolation according to the manufacturer’s protocol, beginning with the RNA bead binding step. RNA concentration and quality were assessed using the Nanodrop 2000 spectrophotometer (Thermo Scientific) and the RNA 6000 Nano LabChip assay (Agilent Technologies).

Libraries were prepared by the Genome Technologies core facility at The Jackson Laboratory using the KAPA Stranded mRNA-Seq Kit (KAPA Biosystems), according to the manufacturer’s instructions. Briefly, the protocol entailed isolation of polyA containing mRNA using oligo-dT magnetic beads, RNA fragmentation, first and second-strand cDNA synthesis, ligation of Illumina-specific adapters containing a unique barcode sequence for each library, and polymerase chain reaction (PCR) amplification. Libraries were checked for quality and concentration using the DNA 1000 LabChip assay (Agilent Technologies) and quantitative PCR (KAPA Biosystems), according to the manufacturer’s instructions. A total of 36 liver samples were collected (3 male and 3 female *Aldh2* knockout mice, 3 male and 3 female *Abca6* knockout mice, and 3 male and 3 female wildtype mice, for each of the two diets). A library was made for each sample followed by pooling of 6 libraries and samples were sequenced by the Genome Technologies core facility at The Jackson Laboratory, 125 bp paired-end on the HiSeq 2500 system (Illumina, Inc.; San Diego, CA) using the TruSeq SBS Kit v4 reagents (Illumina, Inc.) with a minimum of 40M reads per sample. A repeated measures two-way ANOVA test with post-hoc pairwise comparison was used to test for differentially expressed genes between wild type and knock-out mouse. A significant difference was determined at a false discovery rate <0.05 to account multiple testing.

## Data Availability

The genotype data, gene expression, phenotype data that support the findings from the FHS of this 403 study have been deposited in dbGaP (dbGaP Study Accession: phs000363.v16.p10).

## Data availability

The genotype data, gene expression, phenotype data that support the findings from the FHS of this study have been deposited in dbGaP (dbGaP Study Accession: phs000363.v16.p10).

## Supplementary Materials

Fig. S1. The functional analyses of differentially expressed genes generated through IPA.

Fig.S2. Interaction networks of molecules based on known relationships in the QIAGEN Knowledge

Table S1. Clinical Characteristics of the Framingham Heart Study Participants. Table S2. SNPs from Lipids GWAS

Table S3. GWAS SNPs associated with gene expression(eQTLs) in FHS

Table S4. Mediation effect of SNPs on lipids through gene expression

Table S5. CVD proteins and their first-degree interactors from protein-protein interaction network

Table S6. Interactions between CVD proteins

Table S7. Differentially expressed genes from RNA-Seq after ALDH2 and ABCA6 knockout

Table S8. Overlap and molecular functions of differentially expressed genes between *Aldh2* and *Abca6* knockout mice

## Acknowledgments

We gratefully acknowledge the contribution of the Reproductive Science, Histopathology, and Genome Technologies Services at The Jackson Laboratory for expert assistance with the work described in this publication.

## Author contributions

Conceptualization, C.Y. and D.L. Methodology and data analysis, C.Y. Mouse knockout experiments, H.S.,Y.T.,R.K. Protein interaction network, T.H.,W.B.,D.E.H, M.V. Writing, C.Y., R.K.,D.L.

## Data and materials availability

The SNP, gene expression and protein phenotype data that Support the findings from the FHS of this study have been deposited in dbGaP (dbGaP Study Accession: phs000363.v16.p10).

